# Emergence in late 2020 of multiple lineages of SARS-CoV-2 Spike protein variants affecting amino acid position 677

**DOI:** 10.1101/2021.02.12.21251658

**Authors:** Emma B. Hodcroft, Daryl B. Domman, Daniel J. Snyder, Kasopefoluwa Y. Oguntuyo, Maarten Van Diest, Kenneth H. Densmore, Kurt C. Schwalm, Jon Femling, Jennifer L. Carroll, Rona S. Scott, Martha M. Whyte, Michael W. Edwards, Noah C. Hull, Christopher G. Kevil, John A. Vanchiere, Benhur Lee, Darrell L. Dinwiddie, Vaughn S. Cooper, Jeremy P. Kamil

**Affiliations:** Institute of Social and Preventive Medicine, University of Bern, Switzerland; University of New Mexico Health Sciences Center, Albuquerque, NM, USA; Microbial Genome Sequencing Center, LLC, Pittsburgh, PA, USA; Icahn Mt Sinai School of Medicine, New York, New York. USA; Louisiana State University Health Sciences Center, Shreveport, Shreveport, LA, USA; Louisiana Department of Health. Minden, LA, USA; New Mexico Department of Health, Albuquerque, NM, USA; Wyoming Public Health Laboratory, Cheyenne, WY, USA; University of Pittsburgh, School of Medicine, Pittsburgh, PA, USA

## Abstract

The severe acute respiratory syndrome coronavirus 2 (SARS-CoV-2) spike protein (S) plays critical roles in host cell entry. Non-synonymous substitutions affecting S are not uncommon and have become fixed in a number of SARS-CoV-2 lineages. A subset of such mutations enable escape from neutralizing antibodies or are thought to enhance transmission through mechanisms such as increased affinity for the cell entry receptor, angiotensin-converting enzyme 2 (ACE2). Independent genomic surveillance programs based in New Mexico and Louisiana contemporaneously detected the rapid rise of numerous clade 20G (lineage B.1.2) infections carrying a Q677P substitution in S. The variant was first detected in the US on October 23, yet between 01 Dec 2020 and 19 Jan 2021 it rose to represent 27.8% and 11.3% of all SARS-CoV-2 genomes sequenced from Louisiana and New Mexico, respectively. Q677P cases have been detected predominantly in the south central and southwest United States; as of 03 Feb 2021, GISAID data show 499 viral sequences of this variant from the USA. Phylogenetic analyses revealed the independent evolution and spread of at least six distinct Q677H sub-lineages, with first collection dates ranging from mid-August to late November 2020. Four 677H clades from clade 20G (B.1.2), 20A (B.1.234), and 20B (B.1.1.220, and B.1.1.222) each contain roughly 100 or fewer sequenced cases, while a distinct pair of clade 20G clusters are represented by 754 and 298 cases, respectively. Although sampling bias and founder effects may have contributed to the rise of S:677 polymorphic variants, the proximity of this position to the polybasic cleavage site at the S1/S2 boundary are consistent with its potential functional relevance during cell entry, suggesting parallel evolution of a trait that may confer an advantage in spread or transmission. Taken together, our findings demonstrate simultaneous convergent evolution, thus providing an impetus to further evaluate S:677 polymorphisms for effects on proteolytic processing, cell tropism, and transmissibility.

## Introduction

In mid-December 2020, the United Kingdom reported a SARS-CoV-2 variant termed B.1.1.7 (20I/501Y.V1) that exhibited a rapid increase in its range and incidence following its initial detection in November (Andrew Rambaut, Nick Loman, Oliver Pybus, Wendy Barclay, Jeff Barrett, Alesandro Carabelli, Tom Connor, Tom Peacock, David L Robertson, Erik Volz, COVID-19 Genomics Consortium UK (CoG-UK), 2020; Volz, Mishra, *et al*., 2021). Since then, additional “variants of concern” have emerged, namely lineages B.1.351 (20H/501Y.V2) from South Africa (Tegally *et al*., 2020) and P.1 (20J/501Y.V3) and P.2 from Brazil (Voloch *et al*., 2020; Faria *et al*., 2021; Naveca, da Costa, *et al*., 2021; Sabino *et al*., 2021). A key concern is that certain polymorphisms may enhance SARS-CoV-2 infectivity or transmission, akin to what was seen for Spike D614G (Korber *et al*., 2020; Volz, Hill, *et al*., 2021), which has overtaken the original D614 form of the virus that dominated at the outset of the pandemic.

In areas where SARS-CoV-2 seroprevalence is high due to elevated rates of transmission during primary waves of the pandemic, selection pressures may have favored the emergence of variants that escape neutralizing antibodies. Such circumstances are thought to have contributed to the rise of lineages B.1.351 and P.1 (501Y.V2 and 501Y.V3), which in addition to Spike N501Y, harbor at least two other non-synonymous substitutions, K417N/T and E484K, which have been found to confer escape from neutralizing antibodies (Weisblum, Schmidt, Zhang, DaSilva, Poston, J. C. C. Lorenzi, *et al*., 2020; Cele *et al*., 2021; Liu *et al*., 2021; Wang *et al*., 2021).

Despite accounting for roughly 25% of globally recorded COVID-19 cases, and approximately 20% of the available SARS-CoV-2 genome data, relatively few studies have detailed the introduction and emergence of SARS-CoV-2 lineages in the United States (Rochman *et al*., 2020; Worobey *et al*., 2020; Washington *et al*., 2021; Zeller *et al*., 2021). Furthermore, seroprevalence surveys indicate that between 1 in 10 and 1 in 3 people in the USA have already been infected with SARS-CoV-2, potentially high enough that selection for immune evasion may be ongoing (Angulo, Finelli and Swerdlow, 2021; Bubar *et al*., 2021).

Variants affecting the Spike (S) protein are of great interest due to their potential to impact transmissibility or to escape neutralizing antibodies developed in response to natural infection or vaccines. A lineage carrying a D614G substitution quickly came to dominate the pandemic (currently accounting for >98% of sequences)(Korber *et al*., 2020), at least in part because this substitution promotes an S conformation that is more competent for binding to angiotensin-converting enzyme 2 (ACE2) (Yurkovetskiy *et al*., 2020) and reduces shedding of the S1 subunit that contains the receptor binding domain (Zhang *et al*., 2020). Missense mutations at other positions, for example S477N and N439K, have appeared multiple times in large infection clusters in Australia and Europe and are associated with resistance to certain antibodies and/or increased affinity for ACE2 (Hodcroft *et al*., 2020; Liu *et al*., 2020; Weisblum, Schmidt, Zhang, DaSilva, Poston, J. C. Lorenzi, *et al*., 2020; Gaebler *et al*., 2021; Thomson *et al*., 2021).

Here, we describe evidence from two independent SARS-CoV-2 genomic surveillance programs in Louisiana and New Mexico that each detected the rise of S variants affecting position 677 in the later months of 2020. We further provide phylogenetic analyses that identify six independent Q677H sub-lineages and one Q677P sub-lineage that all appear to have emerged within the United States. These variants were not detected until mid-August 2020, but as of 03 Feb 2021 already account for over 2,327 of the 102,462 genomes shared by USA based submitting and collecting authors via the GISAID initiative (Elbe and Buckland-Merrett, 2017; Shu and McCauley, 2017). Given the broad detection of the lineages across multiple states and the apparent increase in their frequencies, these novel emergent Q677H and Q677P lineages merit further study for potential differences in transmissibility.

## Results

In late January of 2021, our two independent SARS-CoV-2 genomic surveillance programs, based at the University of New Mexico Health Sciences (UNM HSC) in Albuquerque, New Mexico and the Louisiana State University Health Sciences Center (LSUHS) in Shreveport, Louisiana, each noticed increasing numbers of PANGO lineage B.1.2 (Rambaut *et al*., 2020) / Nextstrain clade 20G viruses carrying an S:Q677P mutation, and that this variant had increased in frequency in samples collected in late 2020 to mid January (**FIG. 1**). We also noted broad but uneven geographical distribution of these S: 677 polymorphic viruses across the United States, which approached 15% of total viral sequences submitted since the beginning of the pandemic in certain states. Given the proximity of this residue to the polybasic cleavage site (681-685), our observations prompted communication between our two surveillance programs and motivated us to carry out phylogenetic analyses of variation at S position 677.

**Figure 1.**
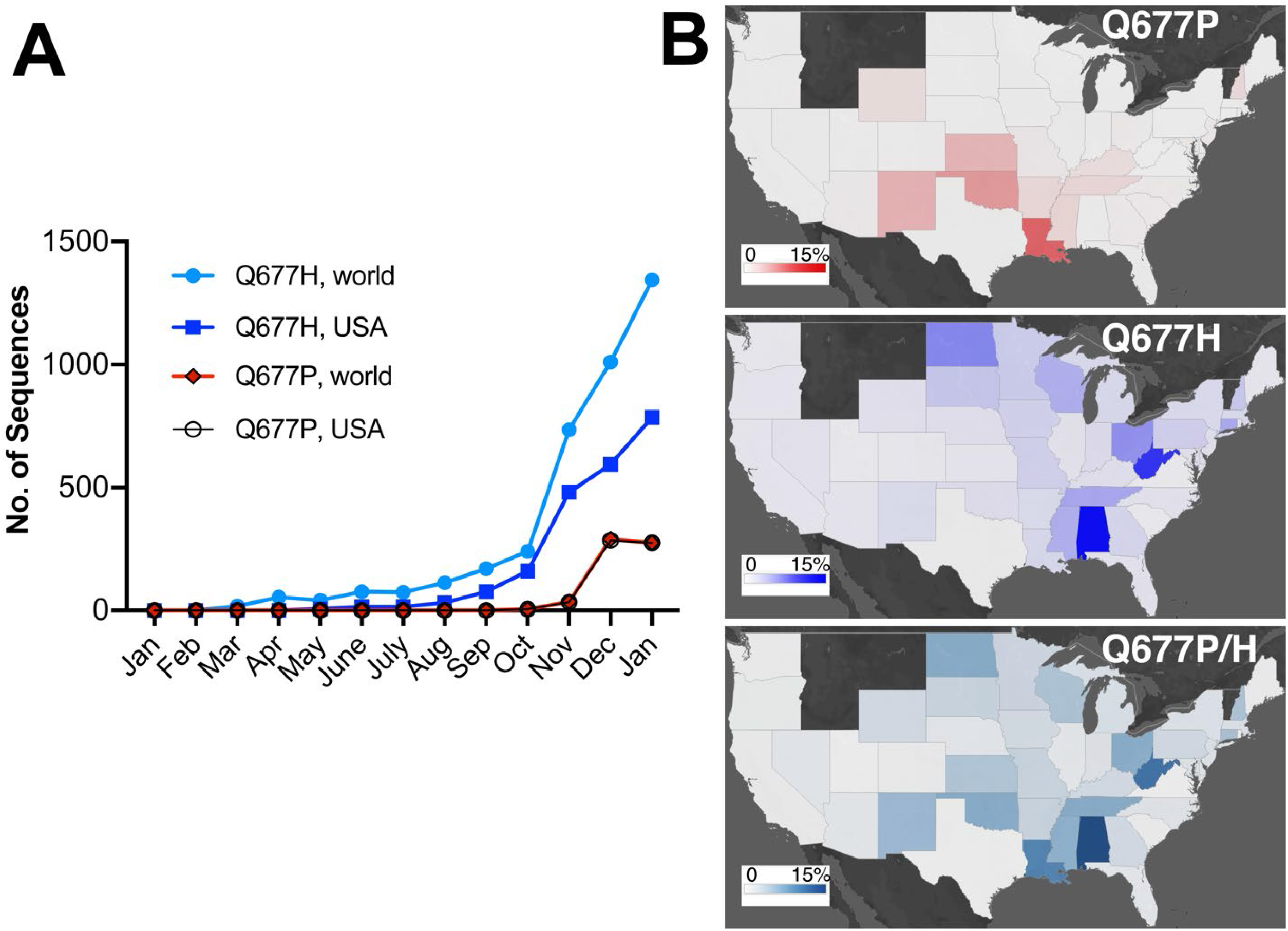
Rising prevalence of SARS-CoV-2 S:Q677P and S:Q677H variants in the United States. (**A**) Monthly number of Q677H and Q677P variant viruses in the USA and worldwide found in GISAID data. Note that data for samples collected in December and January are incomplete for many surveillance facilities. (**B**) State-by-state prevalence of United States Q677H and Q677P variants (percent frequency shaded from 0%-15% out of total GISAID data up to Feb 03, 2021).

### Site frequency dynamics

In addition to the Q677P sub-lineage, our analyses indicated that SARS-CoV-2 variants carrying non-synonymous mutations affecting S codon 677 have arisen at least six other times in the United States (**FIG. 2, FIG. 3**). Four of these occurred within clade 20G, a large clade that has accounted for over 30% of US sequences since October 2020, and as of the beginning of February 2021 represents approximately 50% of all SARS-CoV-2 sequence data in the USA (*Nextstrain Build, North America, Feb 04, 2021*, 2021). The amino acid Q changes to H due to a mutation at nucleotide position 23593. Notably, in four of these six lineages, the mutation changes from G to U, whereas in the other two, it changes from G to C (**FIG. 2**). In contrast, the S:Q677P variant occurs by virtue of an A to C change at position 23592. All mutations leading to Q677H or Q677P involve transversions. Hence, their spontaneous occurrence is generally disfavored relative to transitions. In SARS-CoV-2 samples from human infections, A to C and G to C transversions occur at only ∼10% the frequency of C to U transitions, while G to U mutations are more common, occurring half as frequently as C to U. (Wright, Lakdawala and Cooper, 2020) (Ratcliff and Simmonds, 2021). We also note in GISAID data an S: Q677R cluster of 75 sequences caused by an A to G transition at position 23592 (not shown). However, we focus here on the Q677P and Q677H variants. For ease of discussion and to avoid using geographically-associated names or nicknames, we have named each of the remaining seven S: 677 mutant lineages identified here after American bird species.

**Figure 2.**
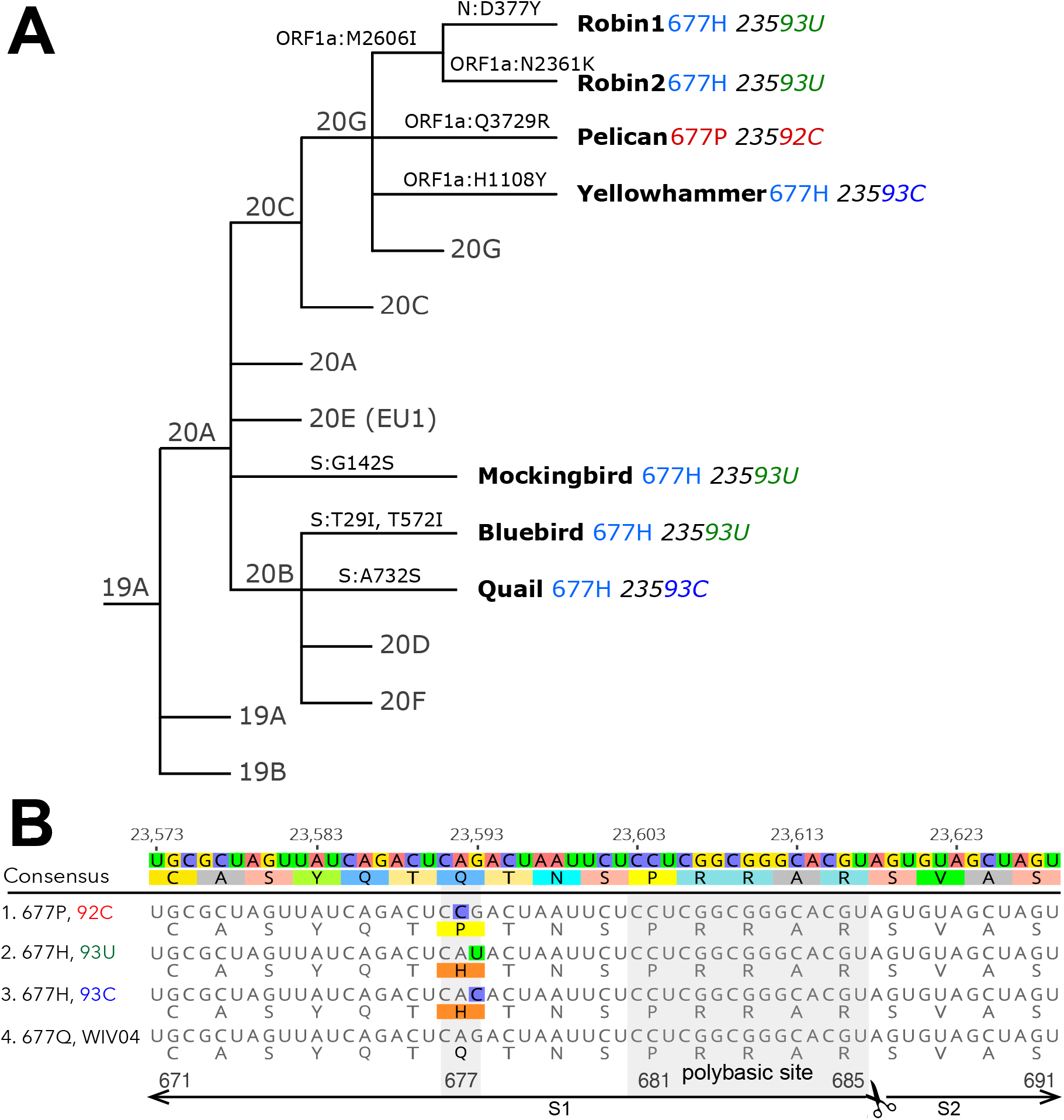
Newly emergent SARS-CoV-2 spike position 677 variants. (**A**) A simplified Nextstrain phylogeny of 677H and 677P lineages. (**B**) A nucleotide and protein alignment of representative S genes from Q677P 92C, and Q677H 93U, Q677H 93C lineages, set against the reference sequence: Dec 2019 Wuhan, China, WIV04 (GISAID: EPI_ISL_402124, GenBank: MN996528). The polybasic or “furin” cleavage site is labeled, as are the portions of the S1 and S2 subunits pictured in the alignment.

**Figure 3.**
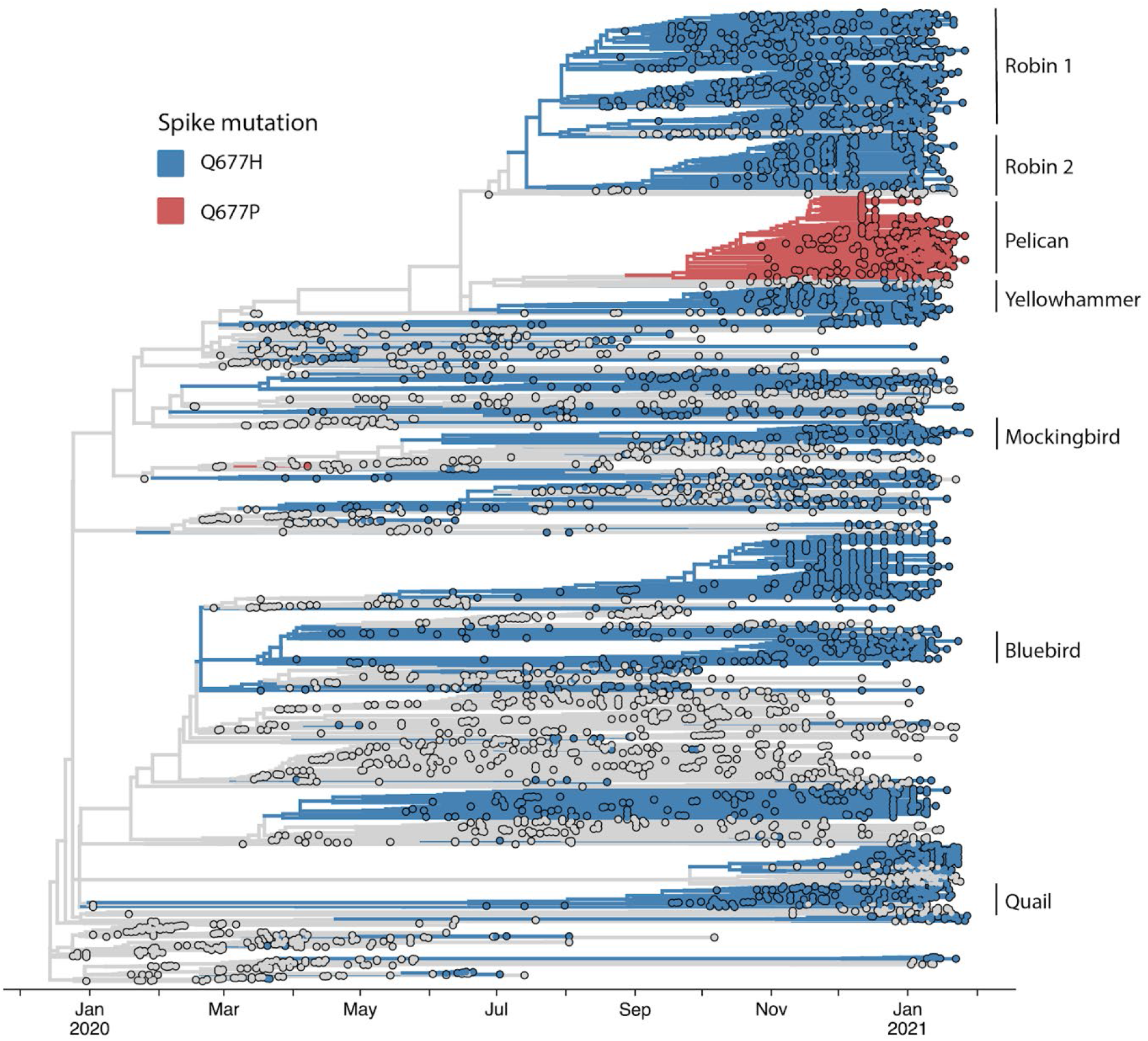
Time-scaled phylogeny of recently expanded United States S:677 variant lineages in their international context. A ‘focal’ Nextstrain build was prepared initially selecting for sequences with any mutation at position 23592 or 23593. These sequences were used as a ‘focal set’ around which background sequences are selected, capturing both the sequences most genetically similar to the focal set, as well as a selection of sequences distributed across time and by country. Red and blue indicate Q677H and Q677P viral genome sequences, as indicated in the legend. USA ‘bird’ lineages are specifically labeled among global data for S:677 variants represented in the build. A full table of acknowledgements for the data included in this analysis is available in the supplement.

### Six U.S. lineages and a provisional naming system

The largest of the 677 variant sub-lineages (“Robin 1”) is a B.1.2 / 20G clade virus carrying Q677H that first appears in GISAID data from a sample with a August 17, 2020 collection date. As of Feb 4, 2021, this sub-lineage contained 754 sequences (**Table 1, FIG. 3**). Robin 1 is found in over 30 US states, but predominates in the Midwest. A second Q677H clade, distinguished from Robin 1 by an N2361K substitution in orf1a, first appeared from a Oct 6, 2020 sample from Alabama and is named “Robin 2” owing to its similarity to the parental Robin 1 sub-lineage. This cluster contains 303 sequences, and is found mostly in the Southeast. Of note, a pre-print study recently reported novel “20C-US” lineage, which includes Robin 1, and made mention of S:Q677P and S:Q677R variants in the US (Pater *et al*., 2021).

**Table 1.** Genetic attributes and relative abundance of six different SARS-CoV-2 lineages identified by S:Q677 mutations.

|  | **667H**<br>**“Robin1”** | **677H**<br>**“Robin2”** | **677P**<br>**“Pelican”** | **677H**<br>**“Yellowhammer**<br>**”** | **677H**<br>**“Bluebird”** | **677H**<br>**“Quail”** | **677H**<br>**“Mockingbird”** |
| --- | --- | --- | --- | --- | --- | --- | --- |
| **Defining mutation** | G235**93T** | G235**93T** | A235**92C** | G235**93C** | G235**93T** | G235**93C** | G235**93T** |
| **Lineage**<br>**(Nextstrain / pangolin)** | 20G / B.1.2 | 20G / B.1.2 | 20G / B.1.2 | 20G / B.1.2 | 20B / B.1.1.220 | 20B / B.1.1.222 | 20A / B.1.234 |
| **Approximate date of origin** | 2020-08-17 | 2020-10-06 | 2020-10-23 | 2020-11-24 | 2020-08-17 | 2020-10-07 | 2020-11-26 |
| **Deposited sequences** | 754 | 298 | 504 | 125 | 122 | 123 | 52 |
| **S** | D614G<br>Q677H | D614G<br>Q677H | D614G<br>Q677P | L5F<br>D614G<br>Q677H | T29I<br>T572I<br>D614G<br>Q677H<br>D936N | D614G<br>Q677H<br>T732S** | G142S<br>E180V<br>D614G<br>Q677H |
| **ORF3a** | Q57H<br>G172V | Q57H<br>G172V | Q57H<br>G172V | Q57H<br>G172V | S58N<br>H78Y |  |  |
| **ORF1a** | T265I<br>M2606I<br>L3352F | T265I<br>N2361K<br>M2606I<br>L3352F | T265I<br>L3352F<br>Q3729R | T265I<br>M2606I<br>L3352F<br>A3454V |  |  | V665I<br>K2059R<br>D2980G |
| **N** | P67S<br>P199L*<br>D377Y | P67S<br>P199L* | P67S<br>P199L*<br>N:M210I<br>(many) | P67S<br>P199L*<br>A376V<br>E378Q | R203K<br>G204R<br>M234I | R203K<br>G204R | S194L<br>T391I |
| **ORF1b** | P314L<br>N1653D<br>R2613C | P314L<br>T1555I (many)<br>N1653D<br>R2613C | P314L<br>N1653D<br>R2613C | P314L<br>N1653D<br>P1666L<br>R2613C | P314L | P314L | P314L |
| **Example sequence near clade root** | USA/WI-UW-2237/2020 | USA/AL-Cullman-JEMT/2020 | USA/TX-HMH-MCoV-19011/2020 | USA/AL-HGSC-JFBB/2020 | USA/CT-Yale-352/2020 | USA/NY-MSHSPSP-PV19649/2020 | USA/TX-HMH-MCoV-18986/2020 |

\* shared on recent common ancestor within 20G / B.1.2
\*\* T732A occurs on a branch prior to the MRCA of Quail, then A732S occurs on a branch inside of Quail, with only 2 sequences inside the cluster not carrying this mutation.

The next largest cluster is the Q677P variant of 20G (B.1.2) (“Pelican”), which was first detected in Oregon from a sample with collection date of Oct 23, 2020 and as of Feb 3, 2021 contains 504 sequences. The Q677P variant has been detected in LA, NM, NC, WY, MA, ID, MI AZ, CA, TX, WI, and MD, and five international sequences (Australia (2), Denmark, Switzerland, India)(Emma B. Hodcroft, 2021). The remaining Q677H sub-lineages each contain around 100 or fewer sequences, and are named: Yellowhammer, detected mostly in the southeast US; Bluebird, mostly in the northeast United States; Quail, mainly in the Southwest and Northeast; and Mockingbird, mainly in the South-central and East coast states (**Table 1, Fig. 3**). A schematic summarizing the key lineage-specific and shared protein polymorphisms of the US S: Q677P and S:Q677H variants is shown in **FIG 4**.

**Figure 4.**
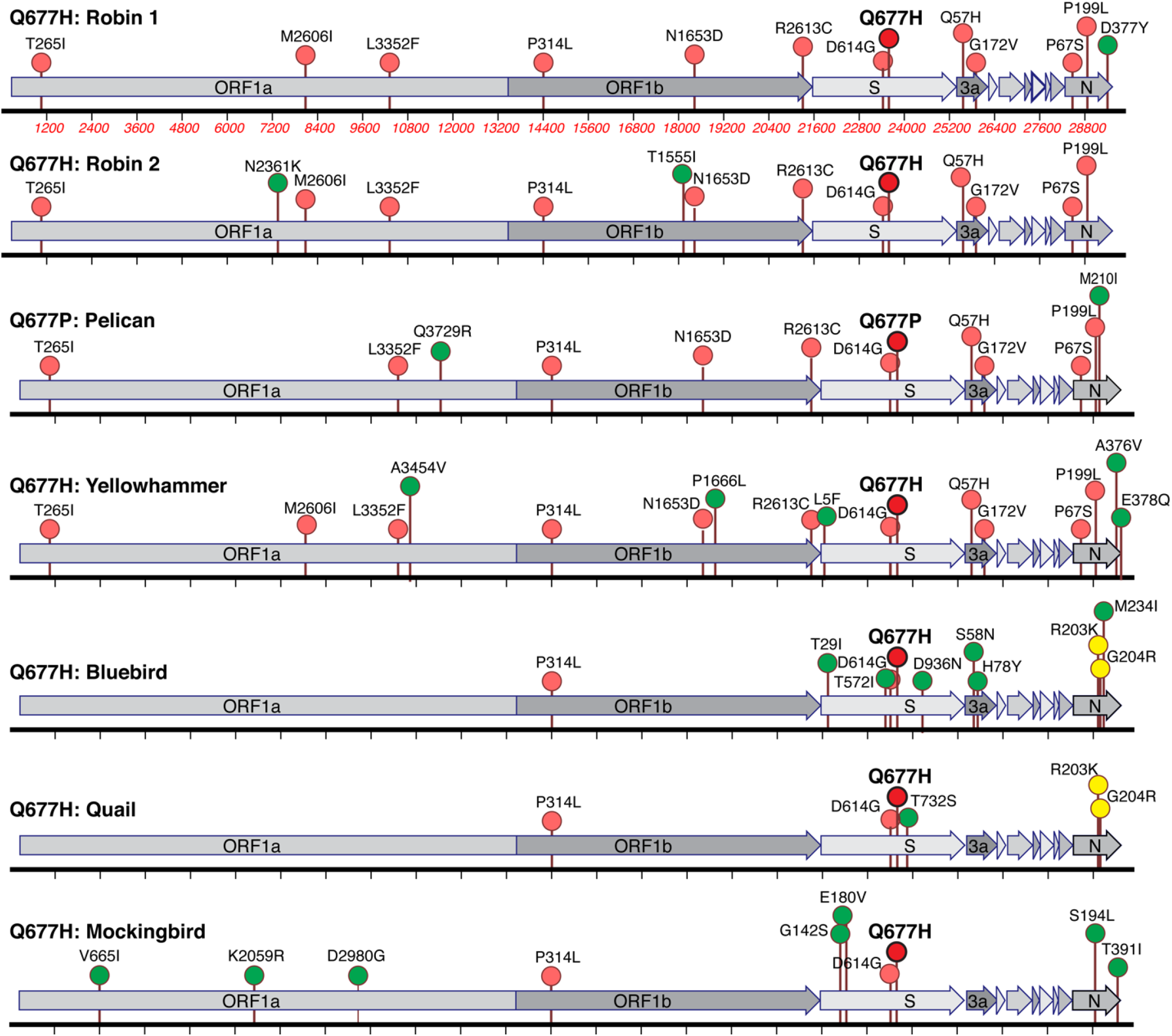
Lineage-specific and shared amino acid polymorphisms of US S:Q677P and S:677H variants. The U.S. S:Q677P and S:Q677H variants defined in Figures 2-3 are illustrated for the major defining protein variants that are shared among most or all of the viruses (pink), or restricted to one (green) or two (yellow). Q677P and Q677H polymorphisms are shown in bold and in red. A full table of acknowledgements for the data included in this analysis is available in the supplement.

### Epidemiological details

Broad-scale genomic surveillance efforts conducted by the New Mexico Department of Health (NM DOH) and the UNM HSC in December and January, 2021 revealed that 83 of 733 SARS-CoV-2 genomes sequenced in New Mexico between December 1^st^ and January 19^th^ contained the S:Q677P variant at a general frequency of 11.3% in the SARS-CoV-2 positive individuals. In New Mexico, the S:Q677P substitution was first observed from a December 12, 2020 sample, and its frequency among sequenced viruses increased through January. In December 2020, our New Mexico-based surveillance effort detected 23 genomes harboring the Q677P mutation, with an additional 59 detected in January 2021. However, limited genomic sequencing of positive cases in New Mexico in October and November hinder the ability to accurately determine the true rate of increase.

Similarly, genomic surveillance efforts conducted by LSUHS together with the Louisiana Department of Health first detected f the S:Q677P variant on Dec 1, 2020, which over the month rose to 187 complete genomes, 150 of which were from a single, large congregate facility outbreak. In part due to deep sampling of the large outbreak, the Q677P virus amounted to 37.2% of all viral genomes sequenced from Dec 2020 samples collected in Louisiana (LSUHS submissions account for 481 of the 503 or 95.6% of the total SARS-CoV-2 genomes available on GISAID for the state with Dec 2020 collection dates). However, the Q677P variant also amounted to 14% of all LSUHS samples collected in Jan 2021, and 11.4% of all Louisiana samples collected between Jan 01 − Jan 19, 2021.

An S:Q677H variant was first detected from Louisiana in the summer of 2020, in LSUHS samples collected on July 21, 2020 and Aug 11, 2020. Only 2 additional S:Q677H sequences were collected from the entire state of Louisiana in Nov, both on the 27^th^. Strikingly, however, the total for Dec 2020 rose to 53, amounting to 10.5% of statewide sequence data for the month. According to the most recent LSUHS data, which covers collection dates up to Jan 19, 2021, the Q677H polymorphism occurs in 5.7% of total SARS-CoV-2 genome data.

When considered in unison, S: Q677P and S: Q677H samples together comprise 47.5% of all Dec 2020 viral genomes collected from Louisiana, and 17.1% of the genome sequences collected in the state from Jan 01 - 19, 2021. Acknowledging that founder effects lead to stochastic fluctuations in the abundance of even neutral genotypes, they also can contribute to the expansion of the fittest viruses. Thus, the observation of a sudden and contemporaneous increase in the abundance of S: Q677H and Q677P variants by surveillance programs in two different states along the southeast / southwest corridor is remarkable.

### Modeled structure of variants

The S1/S2 cleavage site contains the multibasic cleavage site and is found in a disordered region within Spike (S)(Wrapp *et al*., 2020). We used SWISS MODEL to model this inherently flexible region and spotlight proline or histidine residues at position 677 (**FIG. 5**). Although the position 677 is outside the polybasic site (“furin binding pocket”) (Tian, Huajun and Wu, 2012), we speculate that the presence of a proline at this site may introduce a favorable kink that promotes the dynamic conformational changes necessary for cleavage at the S1/S2 junction, which is governed not only by furin-like activities, but also by trypsin-like proteases (e.g., TMPRSS2) and cathepsins (Jaimes, Millet and Whittaker, 2020). Moreover, the introduction of a proline in this model appears to be 3.7 angstroms away from the carbon backbone of S689 (relative to 4.9 angstroms for the native glutamine), which may promote atomic interactions that encourage conformations favorable for proteolytic cleavage. In the case of the S: Q677H substitution, histidine protonation could similarly act as a conformational switch affecting accessibility to proteases. Cleavage at the S1/S2 boundary promotes a more ‘open’ S conformation that is more competent to bind ACE2 (Wrobel *et al*., 2020), and these putative mechanisms for enhanced cleavage at the S1/S2 junction may promote more efficient viral entry (Hoffmann, Kleine-Weber and Pöhlmann, 2020).

**Figure 5.**
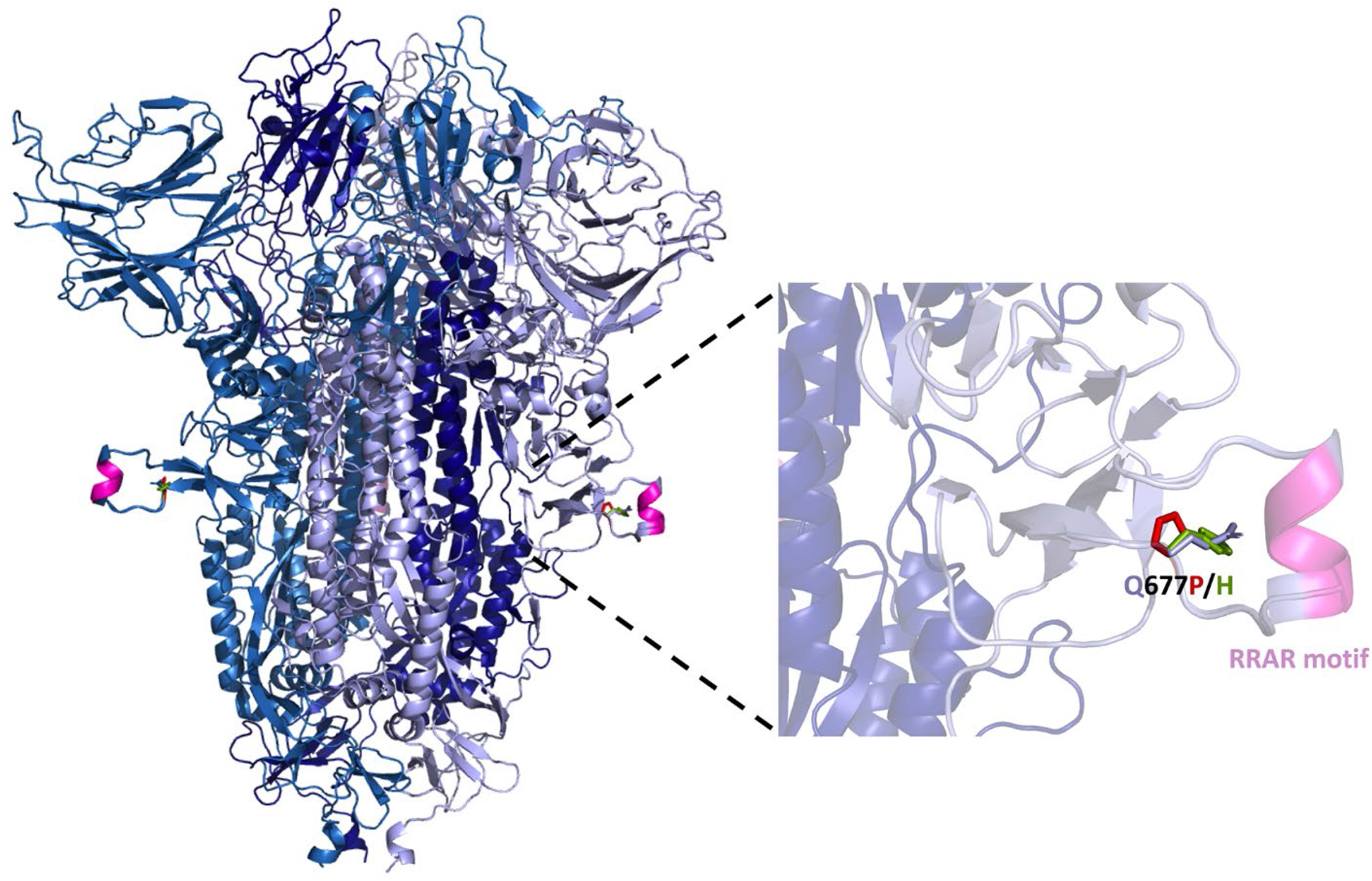
Structure of SARS-CoV-2 Spike protein denoting location of Q677P and Q677H within a disordered loop adjacent to the polybasic (furin) cleavage site. Structure was modeled from PDB: 7BBH using SWISS-MODEL and visualized using PyMol. 677P is shown in red and 677H in green.

## Discussion

### Caveats

Selectively neutral mutations can become fixed in a lineage purely by chance and human behaviour. For instance, the 20E (EU1) lineage characterized by an S: A222V polymorphism emerged suddenly in Europe over the summer, but thus far has not shown evidence for increased transmissibility (Hodcroft *et al*., 2020) and instead is thought to have been spread via holiday travel and relaxing summertime restrictions. Additional S variants such as N439K and S477N also rapidly increased in frequency in Europe over the summer and into the fall of 2020. Although S477N reportedly increases affinity for the entry receptor, ACE2 (Zahradník *et al*., 2021), and both mutations may impact antibody neutralization to some degree (Starr *et al*., 2020; Weisblum, Schmidt, Zhang, DaSilva, Poston, J. C. Lorenzi, *et al*., 2020; Liu *et al*., 2021; Thomson *et al*., 2021), neither shows any signature of increased transmissibility over the S: D614G background from which they emerged, and neither have become prominent in the United States. Therefore, SARS-CoV-2 variants can emerge and increase greatly in number over time in the absence of any clear or sustained selective advantage.

### Convergent evolution is a hallmark of positive selection

The repeated evolution of a trait in independent populations provides strong evidence of adaptation. Between August and November, 2020, seven independent lineages of SARS-CoV-2 with S:Q677H or S:Q677P mutations arose and gained in frequency. This coincidental rise and spread on independent genetic backgrounds is remarkable and suggests some fitness advantage. Observed frequencies undoubtedly incorporate some sampling biases, which may over-estimate the relative amount of S variants affecting position 677. However, sampling bias cuts both ways. U.S. states with fewer deposited sequences may simply have missed detection of these variants. To date, all 677H/P mutants collected from the US stem of the S:614G lineage that now predominates worldwide, but alongside varied polymorphisms in S, N, ORF1a, ORF1b, and other genes, suggesting any fitness advantage of S:677 mutations is largely independent of these other mutations (**Table 1, FIG. 2-4**). Nonetheless, the relatively slow rise of lineages with S:677 substitutions suggest that any fitness benefits may be modest relative to other circulating variants, which may have also independently gained adaptive mutations. Given their relatively recent emergence, however, Q677P/H lineages may continue to rise as a percentage of total cases. Additional laboratory and genetic surveillance data are needed to define whether S: 677 polymorphisms are biologically and clinically relevant.

Although we focus here on the appearance and expansion of S: 677H/P mutants in the United States, global analyses reveal that 677H mutants have arisen multiple times elsewhere in the world as well, including large clusters of 677H mutants in Egypt and Denmark, and multiple clusters in India (Emma B. Hodcroft, 2021). Furthermore, a newly designated, emergent PANGO lineage, B.1.525, carries S: Q677H in addition to several mutations seen in B.1.1.7 (501Y.V1), such as S: del 69-70 and S: del 144, and also, S: E484K (Rambaut *et al*., 2020; Áine O’Toole, 2021). Remarkably, a 19B cluster harboring the ostensibly less fit, ‘ancestral’ D614 Spike, which has been circulating at ≤2% of global frequency since August 2020, recently resurfaced as a newly re-emergent lineage carrying N501Y together with 677H(Wagner, 2021). N501Y is therefore notable for being found in all three “variants of concern” (Tegally *et al*., 2020; *Risk Assessment: Risk related to the spread of new SARS-CoV-2 variants of concern in the EU/EEA – first update*, 2021; Faria *et al*., 2021; Galloway *et al*., 2021; Lauring and Hodcroft, 2021; Naveca, Nascimento, *et al*., 2021; Volz, Mishra, *et al*., 2021). Acknowledging this observation is circumstantial, it further suggests that the 677H mutation may confer an evolutionary advantage to SARS-CoV-2, just as substitutions affecting S positions 417, 484, and 501 have emerged as examples of parallelism in multiple ‘variant of concern’ lineages (Andrew Rambaut, Nick Loman, Oliver Pybus, Wendy Barclay, Jeff Barrett, Alesandro Carabelli, Tom Connor, Tom Peacock, David L Robertson, Erik Volz, COVID-19 Genomics Consortium UK (CoG-UK), 2020; Tegally *et al*., 2020; Naveca, Nascimento, *et al*., 2021; Resende *et al*., 2021).

### The importance of sequencing for viral surveillance and genomic epidemiology

Global surveillance of genomic changes in SARS-CoV-2 varies widely, with leading countries such as Australia, New Zealand, the United Kingdom, and Denmark sequencing viruses from 5-50% of all cases and lagging countries such as the United States, France, Spain, and Brazil sequencing less than 1% of all cases. It is notable that these Q677 variants were detected in the undersampled US population, suggesting that these lineages may actually be more prevalent. The finding of lineages containing 677H mutations in better sampled countries like Denmark indicates that they repeatedly emerge but may be outcompeted by lineages with larger gains in transmission like B.1.1.7 (Davies *et al*., 2020). Collectively, these findings demonstrate the value of greater genomic sequencing and the importance of tracking the emergence and spread of lineages that combine multiple mutations which could enhance transmissibility or evade immunity from prior infection or vaccines.

At least two emergent lineages of concern, B.1.1.7 (501Y.V1), and a newer variant whose prevalence is on the rise in Uganda both contain amino acid substitutions affecting the first position of the polybasic cleavage site, S:P681H and S:P681R, respectively (Andrew Rambaut, Nick Loman, Oliver Pybus, Wendy Barclay, Jeff Barrett, Alesandro Carabelli, Tom Connor, Tom Peacock, David L Robertson, Erik Volz, COVID- 19 Genomics Consortium UK (CoG-UK), 2020; Lule Bugembe *et al*., 2021). The polybasic site strongly impacts viral replication in culture and as well as pathogenesis in animal models (Hoffmann, Kleine-Weber and Pöhlmann, 2020; Johnson *et al*., 2021). Although it is too early to predict whether any particular S: 677 polymorphic lineages will persist, given these observations, the recurrent parallelism affecting S: 677 suggests that this position will continue to surface in variants that show signs of increased transmissibility or fitness. It will be critical to continue genomic surveillance of SARS-CoV-2 to monitor the prevalence of such variants over time, as well as formally define any biological characteristics of these polymorphisms in cell culture and small animal model systems.

## Methods

### Sequencing

Genome sequencing of SARS-CoV-2 at the UNM HSC was conducted from RNA isolated from residual clinical specimens that had previously been determined to be positive for SARS-CoV-2 by FDA-approved diagnostic testing. SARS-CoV-2 genomic sequences were amplified by PCR using the widely adopted ARTIC primer set (v3) and were sequenced either on an Illumina MiSeq or Oxford Nanopore GridION system using in-house, slightly modified versions of protocols (https://www.protocols.io/view/sars-cov-2-illumina-miseq-protocol-v-1-bjd9ki96 & https://www.protocols.io/view/ncov-2019-sequencing-protocol-v3-locost-bh42j8ye), respectively.

For LSUHS samples, SARS-CoV-2 genomic RNA sequencing was done as follows. 13 µL of de-identified total RNA from patient anterior turbinate nasal swabs previously determined as SARS-CoV-2 positive CDC N1 / N2 RT-PCR assay results of Ct <26, was subjected to hybridization capture enrichment sequencing. For each sample, 13µL of extracted RNA was reverse transcribed using Maxima H-minus ds cDNA kits (ThermoFisher Scientific). Libraries were enriched using a Nextera Flex for Enrichment Library Preparation kit with a Respiratory Virus Oligo Set v2, with samples being pooled in 12-plex enrichment reactions, essentially as described elsewhere (O’Flaherty *et al*., 2018). The resulting pools were quantified and grouped in sets of no more than 48 samples and run on a NextSeq 550 using a 150cyc High Output Flow Cell. We used breseq (Deatherage and Barrick, 2014) (v.0.34.1) to map reads to Wuhan-Hu-1 SARS-CoV-2 (NC_045512) or 2019-nCoV WIV04 (GISAID EPI_ISL_402124, NCBI Genbank MN996528)(Zhou et al., 2020) and call the consensus sequence. All predicted mutations were reported for isolates exceeding mean 40x coverage.

### Phylogenetic analyses

A ‘focal’ Nextstrain build of 5874 sequences was prepared as described in Hodcroft et al, 2020, initially selecting for sequences with any mutation at position 23592 or 23593. These sequences are used as a ‘focal set’ around which background sequences are selected, capturing both the sequences most genetically similar to the focal set, as well as a selection of sequences distributed across time and by country. Both of these are then processed through the open-source Nextstrain ‘ncov’ pipeline to produce a time-resolved phylogenetic analysis. A time-stamped version of the Nextstrain build used in this manuscript can be found here: https://nextstrain.org/groups/neherlab/ncov/S.Q677/2020-02-04, and the most recently updated version of this build can be viewed here: https://nextstrain.org/groups/neherlab/ncov/S.Q677. The final designations of the clusters and counts of sequences within them were determined from the resulting phylogenetic tree rather than the ‘raw counts’ from on the nucleotide mutations given above, as the phylogenetic structure can better account for gaps and reversions in sequences which might prevent a sequence being picked up by mutations alone. The resulting JSON file from the Nextstrain pipeline was used to visualize the phylogenetic trees using the baltic package (https://github.com/evogytis/baltic). A full table of acknowledgements for the data included in this analysis is available in the supplement.

### Structural analyses and sequence alignments

SWISS-MODEL(Bienert *et al*., 2017; Waterhouse *et al*., 2018) was used to model the full length, wild type and Q677P mutant spike glycoprotein. The template utilized for the model was 7BBH (Wrobel, A.G., Benton, D.J., Rosenthal, P.B., Gamblin, S.J., 2020), which has all receptor binding domains in the down conformation. The entire furin cleavage site for the D614G (“WT”) and D614G+Q677P (mutant) version of the SARS-CoV-2 Spike, based on PDB. PyMol 2.4 (Schödinger, Inc.) was subsequently used to superimpose the resulting structures and highlight the side chains of each individual structure.

Translation alignment of S:Q677 variants was done using Geneious version 2021.0 (Biomatters Limited, Auckland, NZ).

## Supporting information

Supplemental Table 1 U.S. GISAID entries used in study, by strain

Supplemental Table 2 U.S. GISAID entries used in study, by author

Supplemental Table 3 International GISAID entries used in study, by strain

Supplemental Table 4 International GISAID entries used in study, by author

Supplemental Table 5 All GISAID entries used in study, by strain

Supplemental Table 6 All GISAID entries used in study, by author

## Data Availability

All data for this manuscript is made available via websites and in publicly accessible databases. A time-stamped version of the Nextstrain build used in this manuscript can be found here: https://nextstrain.org/groups/neherlab/ncov/S.Q677/2020-02-04, and the most recently updated version of this build can be viewed here: https://nextstrain.org/groups/neherlab/ncov/S.Q677.

## Acknowledgements

We gratefully acknowledge all submitting authors and collecting authors on whose work this research is based, and to all researchers, clinicians, and public health authorities who make SARS-CoV-2 sequence data available in a timely manner via the GISAID initiative (Elbe and Buckland-Merrett, 2017; Shu and McCauley, 2017).

This work was supported by a COVID-19 Fast Grants award from Emergent Ventures, an initiative of the Mercatus Center at George Mason University (J.P.K.), and by an intramural grant and other funding from the Office of the Vice Chancellor for Research at LSU Health Sciences Center Shreveport (J.P.K., R.S.S., J.A.V.); an Institutional Development Award from the National Institutes of General Medical Sciences of the NIH under grant number P20 GM121307 (C.G.K.); by the Swiss National Science Foundation through grant number 31CA30196046 (E.B.H.), by the U.S. National Center for Research Resources and the National Center for Advancing Translational Sciences of the National Institutes of Health through Grant Number UL1TR001449 (D.L.D., D.B.D), a KL2 Mentored Career Development Award KL2R001448 (D.B.D), and a Translational and Clinical Pilot Project Award CTSC008-11 (D.B.D). We would like to thank the UNM Center for Advanced Research Computing, supported in part by the National Science Foundation, for providing the high performance computing resources used in this work.

## IRB Approvals

SARS-CoV-2 genome sequences were generated under IRB approved protocols STUDY00001445 (LSU Health Sciences Center), and 14-039 and 20-151 (University of New Mexico Health Sciences Center).

## Author contributions

Collected samples: J.A.V., M.W.E., M.M.W, N.C.H.; Sample Preparation: R.S.S., J.L.C.; Generation and processing of sequence data: D.L.D, D.J.S., V.S.C.; Phylogenetic Analyses: E.B.H., D.B.D., V.S.C.; Structural modeling: K.O.; B.L.; Data Curation: K.H.D., C.G.K., M.v.D., D.L.D.; Analysis and Interpretation of Data: E.B.H., D.B.D., D.L.D., M.v.D., K.H.D., C.G.K, K.O., B.L., V.S.C., J.P.K.. Study Design: E.B.H., D.L.D., D.B.D., V.S.C., J.P.K.; Wrote the paper E.B.H., V.S.C., J.P.K., with comments from all authors. Obtained Funding: D.L.D., D.B.D., C.G.K., J.P.K.

## FIGURE LEGENDS

**Supplementary Tables 1, 2, 3, 4, 5, and 6**. Acknowledgement of the Authors and the Originating laboratories where the clinical specimens or virus isolates were first obtained and the Submitting laboratories, where sequence data have been generated and submitted to GISAID. We gratefully acknowledge the Authors from the Originating laboratories responsible for obtaining the specimens and the Submitting laboratories where genetic sequence data were generated and shared via the GISAID Initiative, on which this research is based.

